# Characteristics of Kenyan Women Enrolled in a Trial on Doxycycline Postexposure Prophylaxis for Sexually Transmitted Infection Prevention

**DOI:** 10.1101/2022.04.01.22273292

**Authors:** Kevin Oware, Lydia Adiema, Bernard Rono, Lauren R. Violette, R. Scott McClelland, Deborah Donnell, Caitlin W. Scoville, Josephine Odoyo, Jared M. Baeten, Elizabeth Bukusi, Jenell Stewart

**Affiliations:** Kenya Medical Research Institute (KEMRI), Kisumu, Kenya; Departments of Global Health, University of Washington, Seattle, United States; Medicine (Infectious Diseases), University of Washington, Seattle, United States; Epidemiology, University of Washington, Seattle, United States; Obstetrics and Gynaecology, University of Washington, Seattle, United States; Fred Hutchinson Cancer Research Center, Seattle, United States; Gilead Sciences, Foster City, United States

**Keywords:** STI, cisgender young women, HIV, PrEP, doxycycline post-exposure prophylaxis

## Abstract

**Introduction:** The global incidence of sexually transmitted infections (STIs) has been rapidly increasing over the past decade, with more than one million curable STIs being acquired daily. Young women in sub-Saharan Africa have a high prevalence and incidence of both curable STIs and HIV. The use of doxycycline as a prophylaxis to prevent STI infections is promising; however, clinical trials, to date, have only been conducted among men who have sex with men (MSM) in high-income settings. We describe the characteristics of participants enrolled in the first trial to determine the efficacy of doxycycline post-exposure prophylaxis (PEP) to reduce STI incidence among women.

**Methods:** This is an open-label 1:1 randomized clinical trial of doxycycline PEP efficacy to reduce incident bacterial STIs – *Neisseria gonorrhoeae, Chlamydia trachomatis*, and *Treponema pallidum* – among Kenyan women aged ≥18 and ≤30 years. All were also taking HIV pre-exposure prophylaxis (PrEP). We describe the baseline characteristics of participants.

**Results:** Between February 2020 and November 2021, 449 women were enrolled. The median age was 24 years (IQR 21-27), the majority were never married (66.1%), 370 women (82.4%) reported having a primary sex partner, and 33% had sex with new partners in the 3 months prior to enrolment. Two-thirds (67.5%, 268 women) did not use condoms, 36.7% reported transactional sex, and 43.2% suspected their male partners of having sex with other women. Slightly less than half (45.9%, 206 women) were recently concerned about being exposed to an STI. The prevalence of STIs was 17.9%, with *C. trachomatis* accounting for the majority of infections.

**Conclusion:** Young cisgender women using HIV PrEP in Kenya and enrolled in a trial of doxycycline postexposure prophylaxis had a high prevalence of curable STIs and represent a target population for an STI prevention intervention.

## Introduction

Global trends reveal a rapid increase in the incidence of sexually transmitted infections (STIs) over the past decade, with more than one million curable STIs acquired daily (1). In 2020, the World Health Organization (WHO) estimated 374 million new infections of four curable STIs, *Chlamydia trachomatis, Neisseria gonorrhoeae, Treponema pallidum*, and *Trichomonas vaginalis* (2,3). Young women in sub-Saharan Africa face a high prevalence of curable STIs and HIV (4) and limited data from HIV PrEP trials suggests high incidence rates (5). STIs can severely affect mortality and morbidity for cisgender women by causing conditions such as tubal infertility, chronic pelvic pain, pelvic inflammatory disease, fetal and neonatal death, pregnancy complications, and an increase in susceptibility to HIV (6,7). Women are more biologically predisposed to complications from STIs than men (8). Several studies conducted in sub-Saharan Africa reveal higher STI prevalence among younger women compared to their age-matched male peers and older women (8,9). In the region, the cultural, economic, and social marginalization of women contribute to risk of HIV and STIs (14,15), in part by rendering the negotiation of preventive measures such as condom use, abstinence, and partner notification ineffective (5,10)

Taking antibiotics following a sexual exposure to prevent bacterial STIs places preventive care in the hands of the user. Interventions that are individually controlled are greatly needed, especially for women, and the use of doxycycline as a post-exposure prophylaxis (PEP) has been proposed as a novel STI prevention strategy (11). Doxycycline is already standardly used as prophylaxis to prevent infections such as malaria, Lyme, and leptospirosis (12,13). A recent open-label clinical trial of doxycycline PEP among men who have sex with men (MSM) who were using HIV pre-exposure prophylaxis (PrEP) in France found a 47% relative reduction in bacterial STIs overall and a greater reduction specifically for *C. trachomatis* (70%) and *T. pallidum* (73%) (18). Doxycycline PEP was well-tolerated in that study (18). Several clinical trials of doxycycline as PEP or PrEP among MSM are ongoing worldwide to test this initial finding.

Although women disproportionately bear the burden of adverse sequelae of curable STIs, trials on doxycycline PEP in this population have not yet been completed. The doxycycline Post Exposure Prophylaxis (dPEP) Trial is an open-label, randomized clinical trial evaluating the efficacy of doxycycline PEP for STI prevention (*C. trachomatis, N. gonorrhoeae*, and *T. pallidum*) in Kisumu, Kenya, and is the first study to assess the efficacy of doxycycline PEP in cisgender women. We describe, here, the baseline characteristics of the dPEP Trial population.

## Methods

### Ethics statement

Before implementation, ethical approval of the protocol was received from Kenya Medical Research Institute’s Scientific Ethics Review Unit (KEMRI-SERU) and the University of Washington’s Institutional Review Board. Written informed consent to participate in the study was obtained from all participants before enrolment. The trial is registered with clinicaltrials.gov (#NCT04050540).

### Study design

This is an open-label 1:1 randomized clinical trial evaluating the efficacy of doxycycline PEP to reduce incident curable, bacterial STIs – *N. gonorrhoeae, C. trachomatis*, and *T. pallidum* among Kenyan cisgender young women. Inclusion criteria included willingness and ability to give written informed consent, ≥18 and ≤30 years, female sex assigned at birth, HIV seronegative, and a current prescription for HIV PrEP according to the national guidelines of Kenya. Exclusion criteria included pregnancy, breastfeeding, allergy to tetracyclines, on current medications that may impact doxycycline metabolism or that are contraindicated with doxycycline as per the prescribing information, or recent use of prolonged antibiotics (more than a 14-day course) in the month before enrolment, or active clinically significant medical or psychiatric conditions that would interfere with study participation per the discretion of the study investigator. Participants were primarily recruited from clinics providing HIV PrEP within Kisumu County. Quarterly follow-up visits were scheduled for each participant for 12 months. The study site is situated within a clinic at the Lumumba sub-County hospital in Kisumu, Kenya.

Demographic and behavioral data were electronically collected (REDCap), including questionnaires on STI and HIV risk perception and potential exposures. Biological sample collection (including serum, endocervical and vaginal swab) and testing were done by trained study clinicians and laboratory technologists on site. Rapid HIV testing was completed (Determine) followed by confirmation for any positive results (First Response). Testing for *C. trachomatis* and *N. gonorrhoeae* were done using nucleic acid amplification (Cepheid Xpert or Aptima platforms, depending on availability of testing cartridges). *T. pallidum* was tested using RPR (BD Macro Vue) with TPHA (Fortress Diagnostics) being used to confirm positive results.

This study is an open-label, randomized clinical trial of doxycycline PEP (200mg within 24 hours and up to 72 hours) to reduce bacterial STIs or the standard of care only. Participants were randomized 1:1 to doxycycline PEP vs. standard of care using computer-based randomization (Randomize.net). The trial’s plan to enrol 446 participants was determined based on an anticipated 66 women with new STI infections (N. gonorrhoeae, C. trachomatis, or early syphilis) occurring in the 12 months of follow-up, corresponding to an annual incidence of 22% in the standard of care arm. The trial was designed to achieve 80% power to detect a 50% reduction in infections in the doxycycline arm compared to standard of care. Descriptive analyses were completed using SAS version 9.4 (SAS Institute, Cary, NC, USA).

## Results

Between February 2020 and November 2021, 540 cisgender women were screened for study eligibility and 449 were enrolled. The screening to enrolment ratio was 1.2:1.

**Figure 1:**
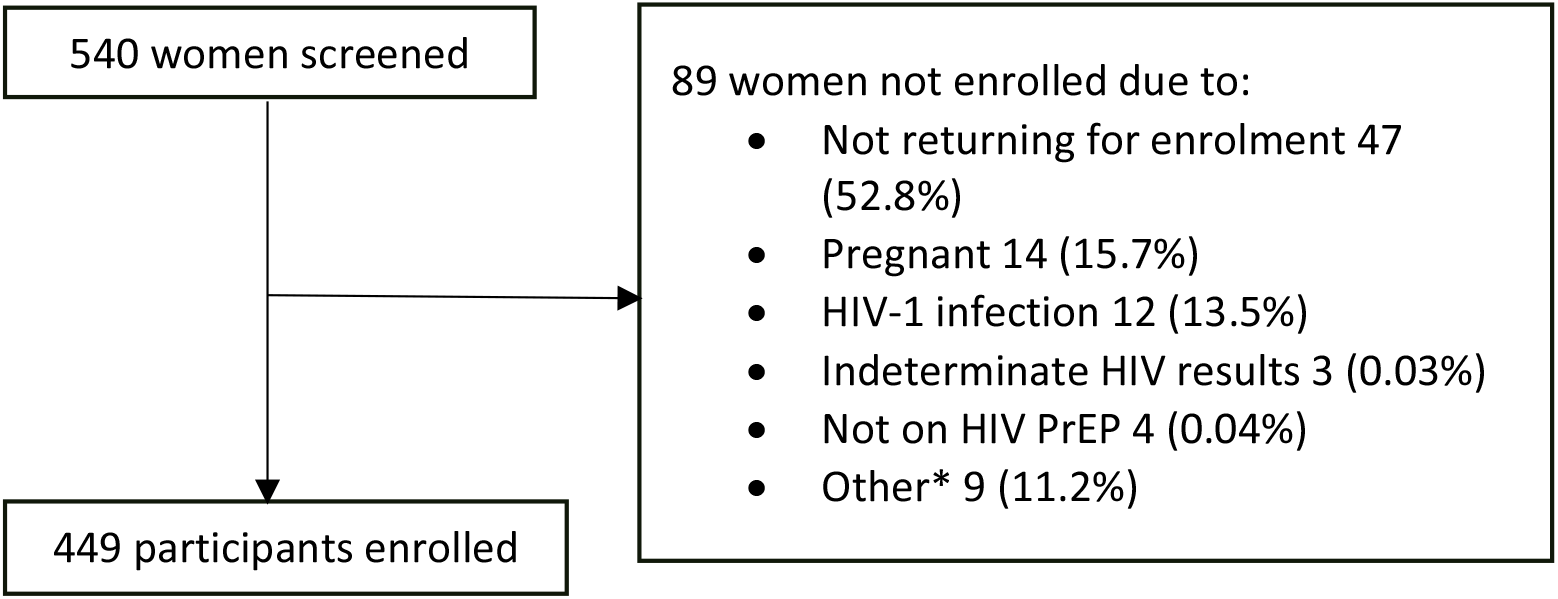
Flow diagram of participant screening and enrolment *Other includes age out of study range, breastfeeding, declined study procedures, comorbidities, and not sexually active.

The median age of participants was 24 years (IQR 21-27) (Table 1). Most were never married (66.1%, 297 women). One hundred and thirty-eight women (30.7%) had not given birth at baseline. Over three quarters (76.8%) had attended at least secondary schooling or higher, and 62.4% (280 women) reported that they earned their own income.

**Table 1.**
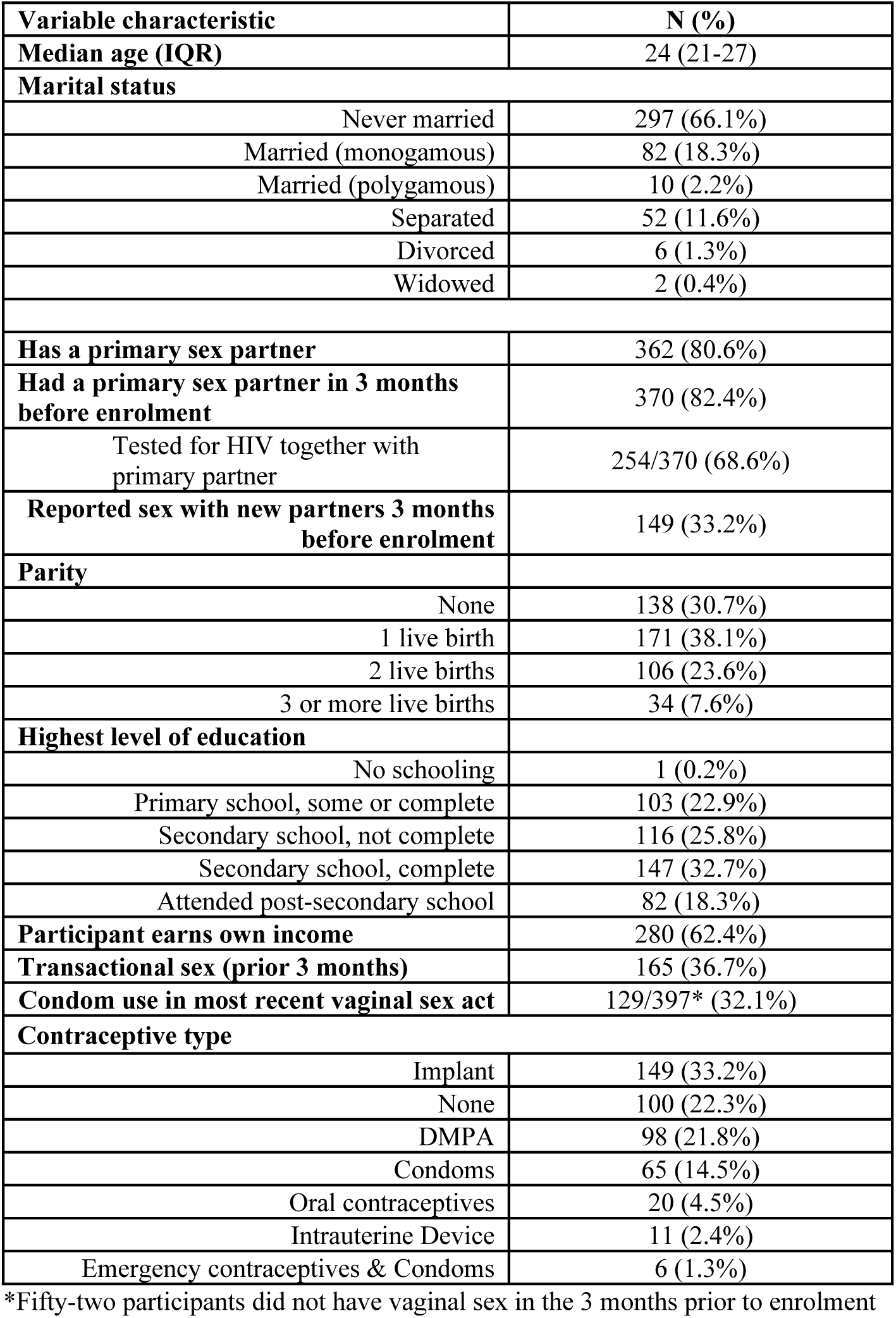
Baseline demographic data, sexual behavior, contraceptive use, and STI prevalence.

The majority of women enrolled (82.4%, 370 women) reported having a primary sex partner in the three months prior to enrolment, and one-third (33.2%, 149 women) reported sex with a new partner in the past three months. A total of 268 (67.5%) women reported not using a condom in the most recent vaginal sex act. Over half (56.6%) had tested for HIV together with their male partners. Recent transactional sex, sex in exchange for goods, gifts, or money, was reported by 36.7% (165 women), and 16.7% (75 women) reported drinking alcohol before sex in the past month.

The majority of women were using long-acting reversible contraceptives: implant 149 (33.2%), injectable depot-medroxyprogesterone acetate 98 (21.8%), and intrauterine device 11 (2.4%). One hundred women (22.3%) reported not using any contraceptive method.

Overall, 17.9% of women (80/448) had any bacterial STI, including 14.1% (63/448) with *C. trachomatis*, 5.8% (17/448) with *N. gonorrhoeae*, and 0.4% (2/449) with *T. pallidum*, where 2 participants (0.4%) presented with both *N. gonorrhoeae* and *C. trachomatis* (Table 2).

**Table 2.**
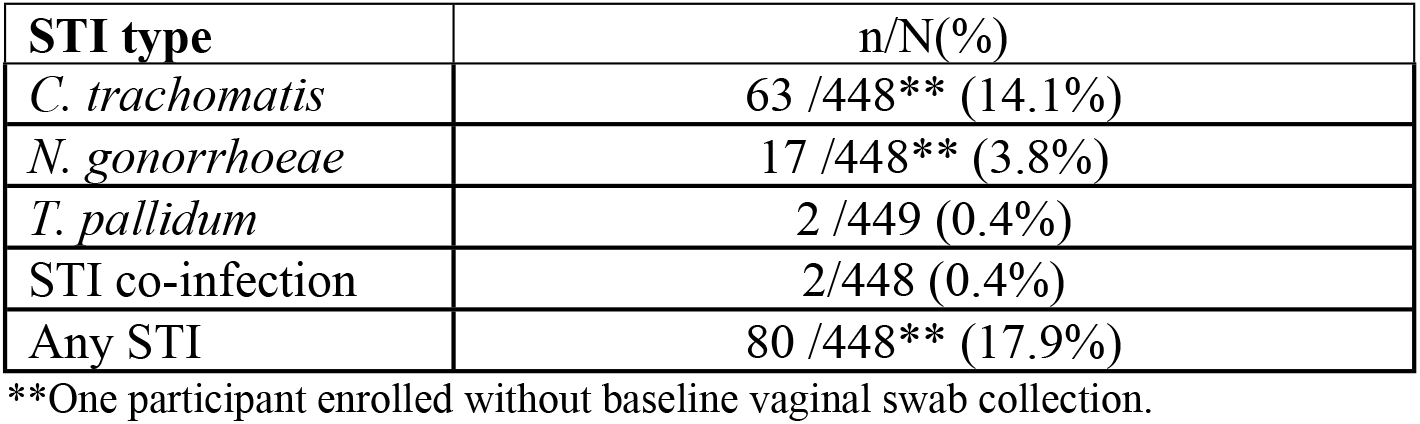
Baseline STI prevalence among cisgender women enrolled in trial of doxycycline postexposure prophylaxis in Kisumu, Kenya.

Participants reported being often or sometimes concerned with contracting STIs (44.5%, 200 women) and HIV (38.1%, 171 women) (Table 3). Cumulatively, about half of the participants (45.9%, 206 women) either agreed or strongly agreed that they were concerned they might engage in sex with someone who could infect them with an STI in the next 3 months. One hundred and eighty women (40.1%) perceived that their sexual behaviour could give them a chance of getting STI in the next 3 months. About half (52.9%) suspected that their male partners might be having sex with someone else.

**Table 3.**
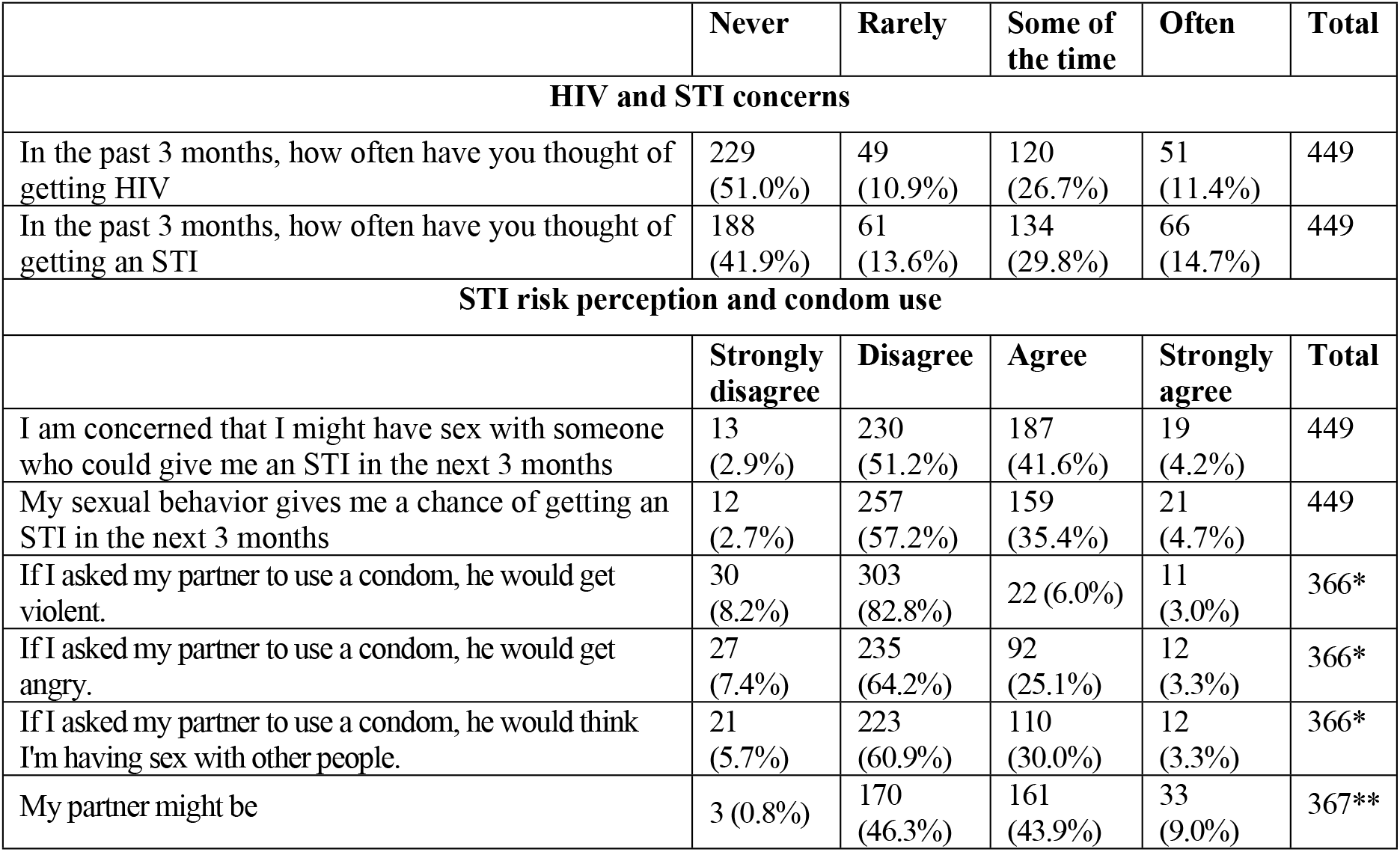
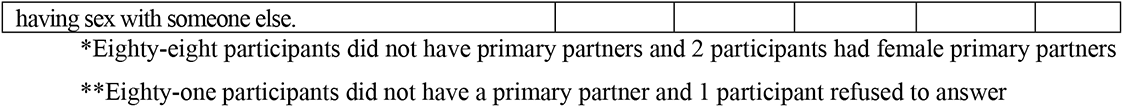
Risk perception among the 449 women taking HIV PrEP and enrolled in the dPEP Trial

## Discussion

The trial enrolled 449 young women who were taking HIV PrEP and half were randomly assigned to take doxycycline PEP. The use of doxycycline as PEP or PrEP for STI prophylaxis is concurrently being evaluated in multiple ongoing trials in the US and Australia among MSM (11,17). The potential for long-term complications that result from a bacterial STI, however, are much greater in cisgender women. The dPEP Trial is among the first to focus on use of doxycycline prophylaxis for primary prevention of bacterial STI among women.

Social structures and sexual behavior that necessitate HIV PrEP use often correlates with a high risk of being exposed to a bacterial STI (4,5). These baseline data indicate that women who are taking HIV PrEP are at significant risk of contracting curable STIs with an overall prevalence of 17.9%, the majority due to *C. trachomatis*. These data support findings from previous studies which equally indicated a high prevalence of *C. trachomatis* among women of reproductive ages in sub-Saharan Africa (9,18). Also consistent with other studies in Kenya, *T. palladium* had the lowest prevalence, with only 2 participants testing positive (19). These baseline data further suggest the need for STI prevention and treatment integrated into PrEP care in sub-Saharan Africa. Furthermore, among the 540 women, primarily recruited from PrEP care, a significant proportion tested positive for HIV at screening (2.2%) highlighting the persistent risk of HIV infection and challenges with adherence to HIV PrEP.

A total of 180 (40.1%) of the cohort identified their own sexual behaviors as possibly increasing risk of STIs with more reporting concerns with the behaviors of their male partners (45.9%). Overall, slightly less than half of the women were concerned about contracting STIs in general. This cohort reported low rates of condom use and fear of conflict with negotiated condom use, highlighting the need for structural interventions beyond individual behavior to reduce the risk of STIs among women (20). Moreover, the low rates of condom use and high rates of STIs in this cohort indicate that women need a strategy to prevent STIs that they can control by themselves since condoms are typically controlled by their male partners.

Health complications that may stem from STI infections among cisgender women include pelvic inflammatory disease, ectopic pregnancy, post-partum endometriosis, and adverse neonatal outcomes like premature death and premature delivery (21). Despite the high frequency of sexual activity, not all study participants reported prior live births, and of the 138 women (30.7%) without prior delivery, 100 (72.5%) were not using hormonal contraception at enrolment. Multiple factors could explain this observation and may be due to frequent use of emergency contraception, induced abortions, miscarriage, and/or infertility. Additionally, a significant number of study participants were using contraceptives, and indicate that women seeking family planning services may benefit from integrating STI prevention, PrEP care, and family planning.

Women of reproductive ages in middle and low-income countries are at increased risk of STI-related complications due to limited access to effective prevention strategies, diagnostic testing, or timely treatment (21,22). The need for primary prevention of STIs is of global importance with highest potential for impact among women in low-resource settings. Overall, young women taking HIV PrEP are at risk of curable STIs and have a demonstrated need for women-centered STI prevention programs.

## Conclusion

Young women using HIV PrEP in Kenya have a high prevalence of bacterial STIs. Should doxycycline PEP be proven to be efficacious at preventing STIs, there is substantial potential for benefit, especially in high prevalence settings such as among women taking HIV PrEP.

## Data Availability

All relevant data are within the manuscript.

## Acknowledgments

This work has been made possible with the acceptance of the young women in Kisumu who volunteered to participate in the study. We thank the study team members (Lawrence Juma, Linda Aswani, Christine Otieno, Violet Kwach, Elizabeth Koyo, Greshon Rota, Loice Okumu, Alfred Obiero) for their contributions to data collection. The Kisumu dPEP Kenya Study staff and Benn Kwach offered valued guidance for manuscript development.

## Author contributions

KO, LA, and JS led the manuscript development, BR and LRV conducted the data analysis. JMB, EB, and JS designed the trial and wrote the protocol. DD was the lead trial statistician. KO, LA, BR, LRV, RSM, DD, CWS, JO, JMB, EB, and JS participated in manuscript preparation.

